# An Adaptive Pragmatic Randomized Controlled Trial of Emergency Department Acupuncture for Acute Musculoskeletal Pain Management

**DOI:** 10.1101/2023.05.16.23290053

**Authors:** Stephanie A. Eucker, Oliver Glass, Mitchell R. Knisely, Amy O’Regan, Alexander Gordee, Cindy Li, Christopher L. Klasson, Olivia TumSuden, Alena Pauley, Harrison Chen, Catherine A. Staton, Maggie Kuchibhatla, Shein-Chung Chow, Duke Emergency Department Acupuncture Research team

## Abstract

**Objective:** Acute musculoskeletal pain in emergency department (ED) patients is frequently severe and challenging to treat with medications alone. The purpose of this study was to determine the feasibility, acceptability and effectiveness of adding ED acupuncture to treat acute episodes of musculoskeletal pain in the neck, back, and/or extremities.

**Methods:** In this pragmatic two-stage adaptive open-label randomized clinical trial, Stage 1 identified whether auricular acupuncture (AA; based on the Battlefield Acupuncture protocol), or peripheral acupuncture (PA; needles in head, neck and extremities only), was more feasible, acceptable and efficacious in the ED. Stage 2 assessed effectiveness of the selected acupuncture intervention(s) on pain reduction compared to usual care only (UC). Licensed acupuncturists delivered AA and PA. They saw and evaluated, but did not deliver acupuncture to, the UC group as an attention control. All participants received usual care from blinded ED providers. The primary outcome was 1-hour change in 11-point pain numeric rating scale.

**Results:** Stage 1 analysis found both acupuncture styles similar. Stage 2 continued all three arms. Among 236 participants randomized, demographics and baseline pain were comparable across groups. The diverse sample recruited was demographically reflective of the larger ED population. Estimated AA+UC (2.1; 95% CI: 1.6, 2.6) and PA+UC (1.6; 95% CI: 1.1, 2.1) 1-hour pain reductions were both significantly greater than UC (0.5; 95% CI: -0.1, 1.0), and participants in both treatment arms reported high satisfaction with acupuncture.

**Conclusion:** ED acupuncture is feasible, acceptable and can effectively reduce acute musculoskeletal pain better than usual care alone.

## INTRODUCTION

### Background

Musculoskeletal disorders are the leading cause of pain and disability, affecting over 1.7 billion people worldwide.^1^ Chronic musculoskeletal pain impairs health, function and well-being, and is associated with chronic opioid use.^1,2^ As chronic pain begins as acute pain and is characterized by acute pain exacerbations, effective acute pain management is essential in mitigating the transition to and/or worsening of chronic pain. Treatment is particularly challenging in emergency department (ED) settings, where acute pain is typically severe, associated with emotional and psychological stressors, and compounded by diagnostic uncertainty.^3,4^ Medications alone provide only a limited degree of pain relief, have potential side effects, and are limited in reducing chronic pain.^5–8^ Moreover, significant disparities exist in the ED treatment of pain, resulting in worse pain outcomes for historically minoritized and underserved populations.^9^ Thus, innovative multi-modal approaches to improve equitable acute musculoskeletal pain management are needed in the ED.

### Importance

International organizations are increasingly calling for nonpharmacologic strategies, including acupuncture, to treat pain.^10,11^ Acupuncture is safe and effective for chronic pain conditions such as cancer pain, neck and low back pain, knee osteoarthritis, and headache.^12,13^ Its analgesic effects are postulated to be mediated through inflammatory and endogenous opioid pathways.^14^ Side effects are typically mild and transient, including needle site pain or bruising, and serious events are extremely rare.^15,16^ While traditional private, hour-long acupuncture sessions are less feasible in space- and time-limited ED environments, more efficient treatment styles have been developed for community clinic and military settings. These include Battlefield Acupuncture, where ear needles are placed in up to 5 specific bilateral auricular acupoints to treat pain,^17,18^ and peripheral acupuncture where needles are placed in acupoints that are easily accessible when fully clothed (e.g. head, neck and extremity acupoints).^15,19^ Recent meta-analyses have shown that collectively the different styles of acupuncture are superior to sham needling^20^ controls (e.g., placing acupuncture needles in non-acupoints, non-penetrating needles, or shallow needles) in the ED and other settings.^21,22^ These studies have also demonstrated physiologic effects with needles beyond placebo, prompting experts to recommend the use of non-sham controls in larger pragmatic trials to assess treatment effectiveness of acupuncture.^23^

Currently, there is limited data on the efficacy of ED acupuncture for acute pain, particularly for diverse and underrepresented populations, and the acupuncture style best-suited to the ED environment remains unclear.^22^ Previous work has been mostly limited to observational or small pilot randomized studies in the U.S. and a few larger trials in other countries.^21,24–26^ Moreover, no study has compared different acupuncture protocols (e.g., battlefield/auricular acupuncture and peripheral acupuncture) to determine which is more feasible, acceptable, or effective in the ED.

### Goals of This Investigation

Therefore, we conducted a pragmatic adaptive randomized control trial of ED acupuncture for acute musculoskeletal pain. A major goal of this study was to expand the indications for acupuncture from prior research to be more generalizable to an ED population experiencing mixed pain conditions (e.g., new acute pain and acute exacerbations of chronic pain). Moreover, we adapted features from two styles of acupuncture to determine which may be more suitable and efficacious in an ED setting: auricular acupuncture (AA) based on the Battlefield Acupuncture protocol, and peripheral acupuncture (PA) based on community acupuncture but restricting needle sites to accessible head, neck and extremity acupoints to treat pain. Thus, the objectives of this study were to: (1) identify which style of acupuncture is feasible, acceptable, and more efficacious for treating acute episodes of musculoskeletal pain in the ED, and (2) determine the effectiveness of that acupuncture style compared to control (no acupuncture) on pain reduction in the ED.

## METHODS AND ANALYSIS

### Study design and setting

This pragmatic, two-stage adaptive open-label randomized clinical trial was approved by the Institutional Review Board (Protocol # Pro00104140), registered on 2/7/2020 with clinicaltrials.gov (registration # NCT04290741) and released to the public on 2/28/2020. The detailed study methods were previously published.^27^ In brief, the first stage objective was to identify which style of ED-based acupuncture, AA or PA, is more feasible, acceptable and potentially more efficacious in ED patients with acute musculoskeletal pain. The second stage objective was to determine the effectiveness of the selected acupuncture style compared to attention control receiving usual care only (UC) on pain reduction. The adaptive approach required a planned interim analysis at the end of Stage 1 to determine whether to make adaptations, such as dropping one of the acupuncture treatment arms for Stage 2, based on the FDA-recommended criterion of probability of being the best treatment.^28,29^ This study took place in a U.S. academic tertiary-care ED with 80,000 visits per year staffed by attending physicians, resident physicians, and physician assistants. We used the CONSORT checklist to report our findings.^30^

### Selection of Participants

Trained clinical research coordinators (CRCs) performed all patient screening, recruitment, informed e-consent, and enrollment procedures. Licensed acupuncturists trained in traditional Chinese medicine explained and delivered acupuncture treatments^27^. A convenience sample of patients was enrolled during 8-hour periods, typically occurring sometime between 8am-8pm, Monday-Friday. English-speaking adult ED patients with acute (≤7 days) new onset or exacerbations of chronic pain in the neck, back, arms and/or legs, deemed musculoskeletal by ED clinicians, were included.^31^ Exclusion criteria included: (1) no pain at triage; (2) contraindication to needles at acupuncture sites (e.g. skin infection); (3) unable to attend clinic; or (4) serious medical condition (e.g., active COVID-19 infection).

### Interventions

Two different acupuncture interventions (AA and PA) were performed by licensed acupuncturists while in the ED, and are described in detail elsewhere.^27^ In brief, (1) <u>Auricular</u> <u>acupuncture (AA)</u> placed press needles in up to 5 bilateral ear sites corresponding to the Battlefield Acupuncture protocol.^17,18^ (2) <u>Peripheral acupuncture (PA)</u> placed needles in select head, neck, and extremity sites based on acupuncturist clinical discretion.^15,19^ (3) As an attention control to account for placebo effects from seeing an additional healthcare provider, the usual care only (UC) participants received the same initial evaluation and pre-treatment interaction with study acupuncturists prior to randomization as the two acupuncture groups, but did not receive acupuncture treatment. All participants in all 3 study arms received usual care at the discretion of their blinded ED clinical team, which in the ED setting typically consists of nonopioid and occasionally opioid medications, less frequently ice or heat packs. These treatments are initially ordered and administered in triage while the patients are in the waiting room after a triage physician assessment, with additional treatments determined by the primary ED team once they were assigned to an ED room.

Participants were randomized 1:1:1 in Stage 1 and 2:2:1 in Stage 2 to AA+UC, PA+UC or UC using a computer-generated unstratified block randomization sequence stored in a secure electronic file, with sequence electronically hidden and visible only at randomization to the acupuncturists. While participants and acupuncturists were not blinded, reasonable attempts were made to blind all other research and clinical personnel, described in detail elsewhere.^22^

### Measurements

Data was collected at ED baseline and at 1 hour post-treatment in a secure REDCap database via iPad in the ED.^32^ Acupuncture treatment details based on Revised Standards for Reporting Interventions in Clinical Trials of Acupuncture (STRICTA) recommendations,^33^ ED medications, and adverse events were recorded by study personnel in REDCap.

### Outcomes

The primary endpoints for Stage 1 were feasibility, acceptability, and safety. Feasibility was assessed by patient recruitment and retention rates, with a goal average of ≥1 patient enrolled and completed 1 hour follow-up per study day. Acceptability was assessed by patient-reported satisfaction with acupuncture treatment, with a goal average of ≥4 on a 5-point Likert scale (from 5=very much to 1=not at all). Safety was evaluated by adverse events (AEs), most commonly non-serious bleeding, bruising, or pain at needle sites, which has been reported in the literature at a rate of roughly 7%.^15,34^ Potentially serious adverse events (SAEs) such as pneumothorax are exceedingly rare (<0.01%).^15,34^ The primary endpoint for Stage 2 was change in current pain score (0-10 numeric rating scale, NRS) from ED baseline to 1-hour post-treatment. We used a minimally clinically significant difference in pain score of 1.3 previously validated in ED patients.^35^

Secondary measures included satisfaction with treatment (1-5 Likert scale) and medication use, including opioid and non-opioid medications, assessed by patient report and electronic medical record (EMR) data. Additional patient self-reported data included demographics, pain characteristics, non-medical opioid use using the Alcohol, Smoking and Substance Involvement Screening Test (ASSIST),^36^ the simplified graded chronic pain scale,^37,38^ as well as pain interference, sleep disturbance, and physical function using the validated Patient-Reported Outcomes Measurement Information System (PROMIS)-29 instruments.^39^ The question timeframe was modified from “over the past 7 days” to “over the past 24 hours (1 day)” for ED baseline assessments due to the acute duration (≤7 days) of pain.

### Analysis

A sample size of 220 total subjects for Phase 1 was calculated using a minimally clinically significant difference in pain score of 1.3, 90% power and α = 5%, based on a 2-stage adaptive design with 90 subjects allocated 1:1:1 to three arms (AA+UC, PA+UC, UC) in Stage 1, and the remaining 130 subjects to two arms assuming 1:2 control:treatment allocation for Stage 2,^29,40,41^ adjusted for one planned interim analysis using O’Brien-Fleming type of alpha spending function^28^ and a 10% drop rate.^27^

Primary analysis for the primary end-point was based on intention-to-treat (ITT) including all randomized subjects combined from both stages with at least one follow-up evaluation. For all outcomes, complete case analysis was conducted by excluding subjects missing the one-hour outcome.

Because usual care for pain in this ED site starts immediately after triage, an exploratory analysis to assess within-group differences in response to usual care vs acupuncture examined differences in pain scores over time from triage to study baseline to 1 hour post-treatment across treatment groups using ANOVA, with the outcome being the difference-of-differences [(one-hour – baseline) – (baseline – triage)].

To determine whether the subgroup of patients with pre-existing chronic pain had a different response to acupuncture than those without chronic pain, we fit an ANOVA model with chronic pain (defined as Grade 2 or 3 on the Graded Chronic Pain Scale) and the interaction of chronic pain and study arm.

All analyses were performed using SAS software version 9.4 (SAS Institute Inc., Cary, NC). Unless otherwise noted, tests of hypotheses were two-sided with 5% level of significance.

### Handling of Missing Data

For the primary ITT analysis, 10 patients were missing change in 1-hour pain score, due to 1 missing baseline and 9 missing 1-hour pain scores. An additional 2 patients recorded 0 pain at baseline (time of enrollment); as all subjects reported pain as their reason for ED visit, these were deemed to be likely data-entry errors and were treated as missing. As NRS pain score was also recorded during the pre-treatment period just prior to randomization, the 3 missing baseline pain scores were imputed using the last observation carried backwards (LOCB) method with the pain score just prior to randomization as the baseline pain value. Multiple imputation methodology was used to impute missing values of the primary outcome of one-hour pain score, wherein the imputation regression model included baseline pain, age, sex, race, and ethnicity as predictors, along with all the first-order interactions of those terms. Change scores were then calculated as estimated reductions from the recorded baseline and imputed one-hour values. Five imputed datasets were generated, and the results of analysis on each of these combined for a final 1-hour change in pain score using standard methodology. For secondary outcomes, missing values were not imputed.

### Analyses

As there were no significant differences in baseline characteristics between groups, the primary outcome was compared using unadjusted ANOVA. Secondary outcomes were compared using separate unadjusted ANOVAs for continuous variables, Kruskal-Wallis for ranked (Likert-scale) variables, and chi-squared for categorical variables. AEs were tabulated, ordered by frequency, and summarized by seriousness, severity, and possible association with acupuncture. Incidence rates of AEs were compared by Fisher’s Exact Test.

### Interim Analysis

One interim analysis was planned at the end of Stage 1 to assess feasibility based on patient recruitment and retention rates, safety based on AEs, and probability of being the more effective treatment arm analyzed from the 1-hour change in pain scores.^29^ An independent data-safety monitoring committee (DSMC), comprised of a biostatistician, emergency physician-researcher and medical acupuncturist, monitored trial safety and performance and recommended adaptations based on interim analysis.

## RESULTS

At interim analysis, one acupuncture arm was not clearly superior to the other, and both trended toward superior compared to control. Therefore, all three study arms were continued in Stage 2 with new allocation ratio 2:2:1 AA+UC:PA+UC:UC.

### Characteristics of Study Subjects

From February 10, 2020 to May 3, 2021, 911 patients were screened, and 236 patients were randomized to one of three arms (68 UC, 84 AA+UC, 84 PA+UC, **Figure 1**). We attained our feasibility goal with on average more than one subject enrolled per day and >95% of subjects in the ITT population completing the one-hour outcome. The study population consisted of broad demographic characteristics representative of the typical ED population seeking care for musculoskeletal pain,^3^ and were similar across the AA+UC, PA+UC and UC arms (**Table 1**). Enrolled subjects had a mean age of 46.1 years (16.5 SD, range: 19 – 85). The most common self-identified race was Black (53.6%), and 7.2% identified as Hispanic. In addition, 56.6% of subjects were employed either full-time or part-time, whereas 21.7% reported being unemployed or unable to work.

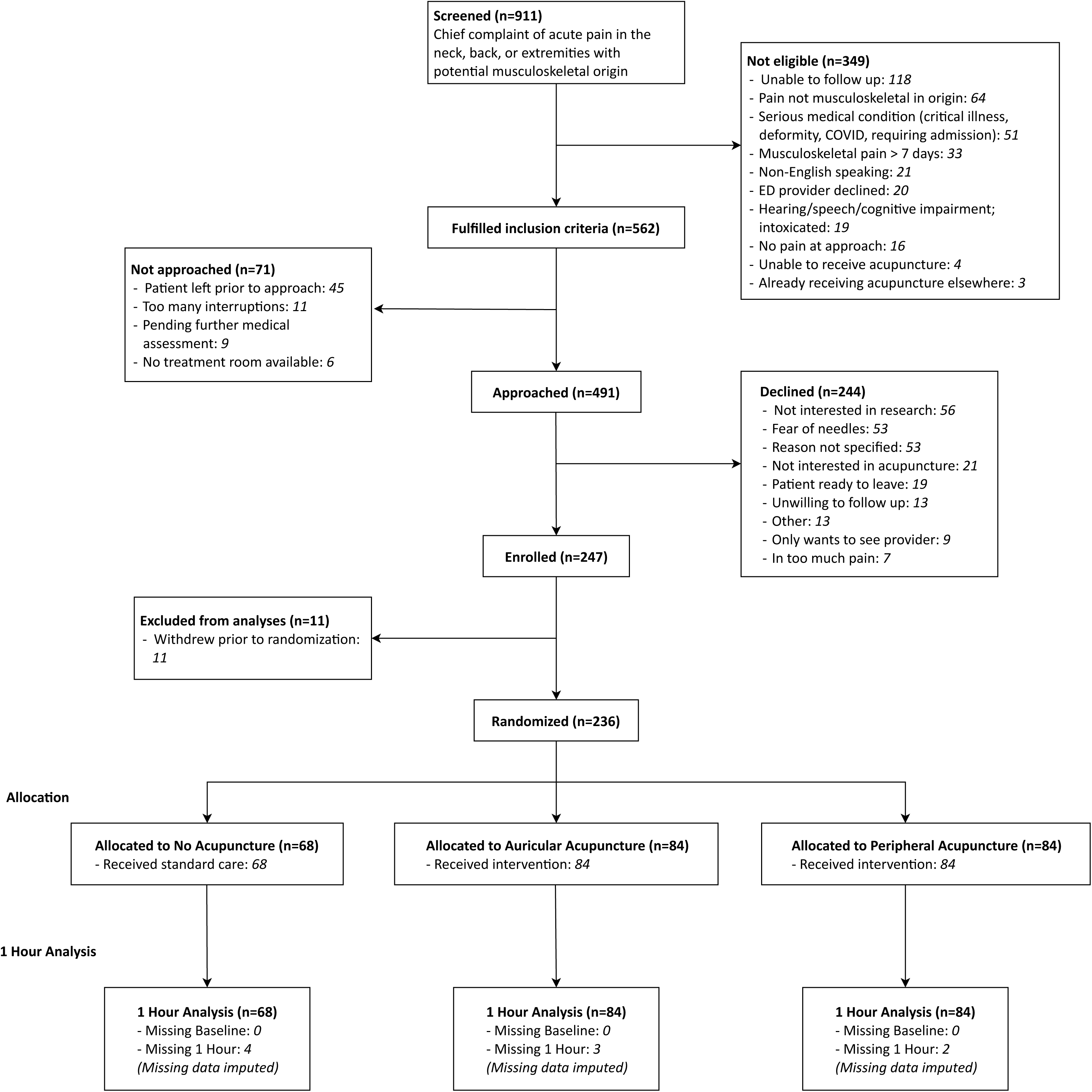

**Table 1:**
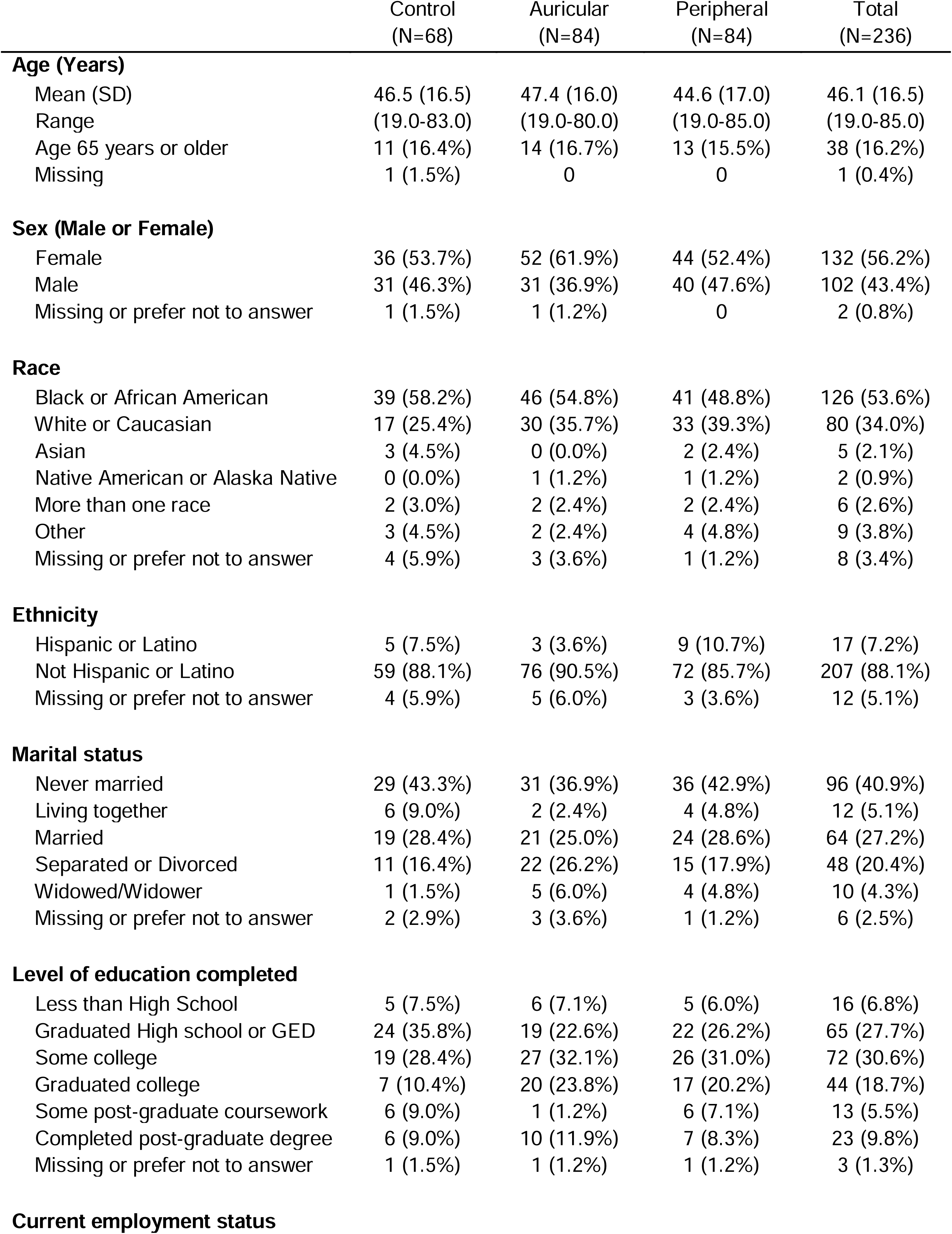
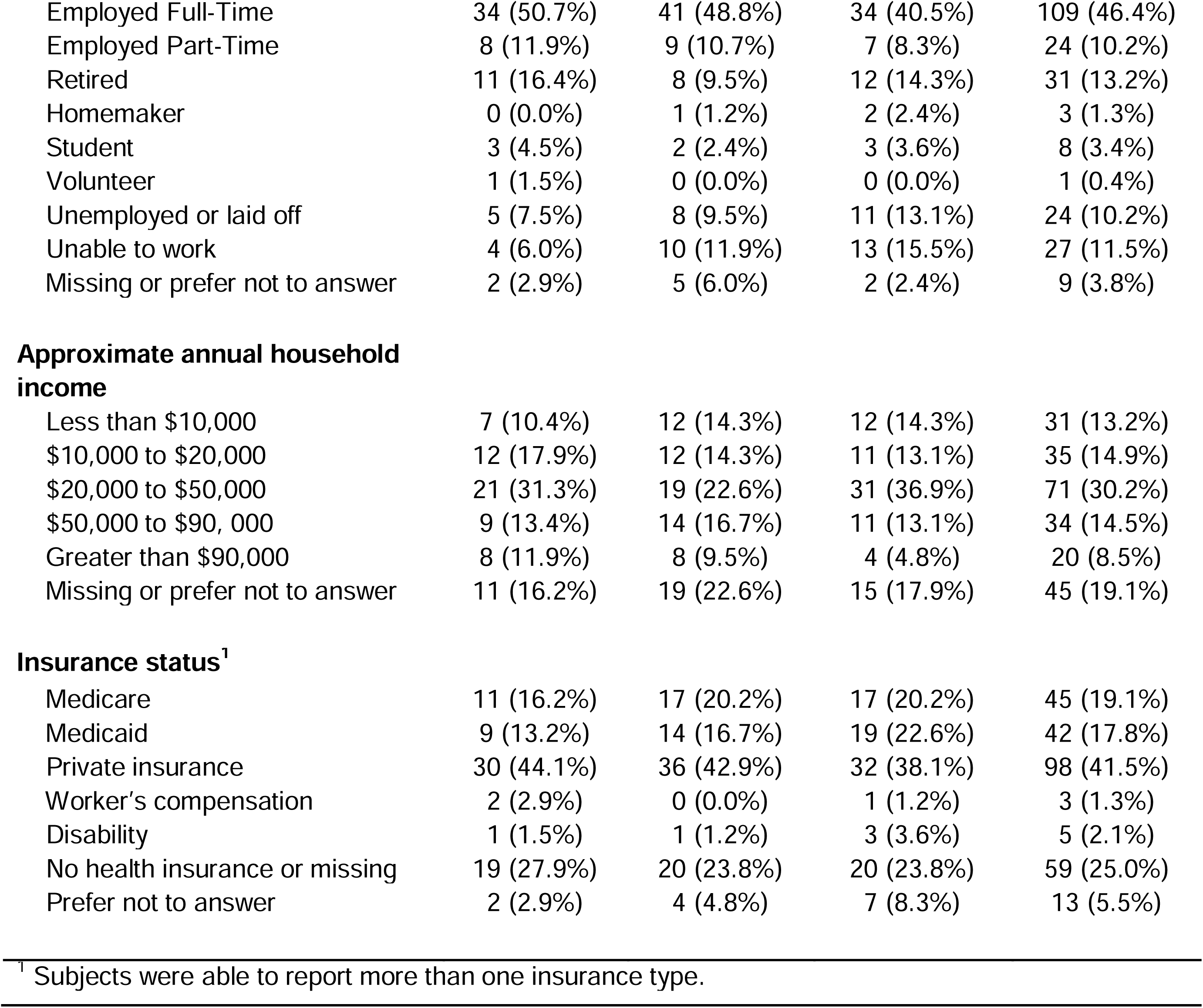
Patient Characteristics.

|  | Control<br>(N=68) | Auricular<br>(N=84) | Peripheral<br>(N=84) | Total<br>(N=236) |
| --- | --- | --- | --- | --- |
| **Age (Years)** |  |  |  |  |
| Mean (SD) | 46.5 (16.5) | 47.4 (16.0) | 44.6 (17.0) | 46.1 (16.5) |
| Range | (19.0-83.0) | (19.0-80.0) | (19.0-85.0) | (19.0-85.0) |
| Age 65 years or older | 11 (16.4%) | 14 (16.7%) | 13 (15.5%) | 38 (16.2%) |
| Missing | 1 (1.5%) | 0 | 0 | 1 (0.4%) |
| **Sex (Male or Female)** |  |  |  |  |
| Female | 36 (53.7%) | 52 (61.9%) | 44 (52.4%) | 132 (56.2%) |
| Male | 31 (46.3%) | 31 (36.9%) | 40 (47.6%) | 102 (43.4%) |
| Missing or prefer not to answer | 1 (1.5%) | 1 (1.2%) | 0 | 2 (0.8%) |
| **Race** |  |  |  |  |
| Black or African American | 39 (58.2%) | 46 (54.8%) | 41 (48.8%) | 126 (53.6%) |
| White or Caucasian | 17 (25.4%) | 30 (35.7%) | 33 (39.3%) | 80 (34.0%) |
| Asian | 3 (4.5%) | 0 (0.0%) | 2 (2.4%) | 5 (2.1%) |
| Native American or Alaska Native | 0 (0.0%) | 1 (1.2%) | 1 (1.2%) | 2 (0.9%) |
| More than one race | 2 (3.0%) | 2 (2.4%) | 2 (2.4%) | 6 (2.6%) |
| Other | 3 (4.5%) | 2 (2.4%) | 4 (4.8%) | 9 (3.8%) |
| Missing or prefer not to answer | 4 (5.9%) | 3 (3.6%) | 1 (1.2%) | 8 (3.4%) |
| **Ethnicity** |  |  |  |  |
| Hispanic or Latino | 5 (7.5%) | 3 (3.6%) | 9 (10.7%) | 17 (7.2%) |
| Not Hispanic or Latino | 59 (88.1%) | 76 (90.5%) | 72 (85.7%) | 207 (88.1%) |
| Missing or prefer not to answer | 4 (5.9%) | 5 (6.0%) | 3 (3.6%) | 12 (5.1%) |
| **Marital status** |  |  |  |  |
| Never married | 29 (43.3%) | 31 (36.9%) | 36 (42.9%) | 96 (40.9%) |
| Living together | 6 (9.0%) | 2 (2.4%) | 4 (4.8%) | 12 (5.1%) |
| Married | 19 (28.4%) | 21 (25.0%) | 24 (28.6%) | 64 (27.2%) |
| Separated or Divorced | 11 (16.4%) | 22 (26.2%) | 15 (17.9%) | 48 (20.4%) |
| Widowed/Widower | 1 (1.5%) | 5 (6.0%) | 4 (4.8%) | 10 (4.3%) |
| Missing or prefer not to answer | 2 (2.9%) | 3 (3.6%) | 1 (1.2%) | 6 (2.5%) |
| **Level of education completed** |  |  |  |  |
| Less than High School | 5 (7.5%) | 6 (7.1%) | 5 (6.0%) | 16 (6.8%) |
| Graduated High school or GED | 24 (35.8%) | 19 (22.6%) | 22 (26.2%) | 65 (27.7%) |
| Some college | 19 (28.4%) | 27 (32.1%) | 26 (31.0%) | 72 (30.6%) |
| Graduated college | 7 (10.4%) | 20 (23.8%) | 17 (20.2%) | 44 (18.7%) |
| Some post-graduate coursework | 6 (9.0%) | 1 (1.2%) | 6 (7.1%) | 13 (5.5%) |
| Completed post-graduate degree | 6 (9.0%) | 10 (11.9%) | 7 (8.3%) | 23 (9.8%) |
| Missing or prefer not to answer | 1 (1.5%) | 1 (1.2%) | 1 (1.2%) | 3 (1.3%) |
| **Current employment status** |  |  |  |  |
| Employed Full-Time | 34 (50.7%) | 41 (48.8%) | 34 (40.5%) | 109 (46.4%) |
| Employed Part-Time | 8 (11.9%) | 9 (10.7%) | 7 (8.3%) | 24 (10.2%) |
| Retired | 11 (16.4%) | 8 (9.5%) | 12 (14.3%) | 31 (13.2%) |
| Homemaker | 0 (0.0%) | 1 (1.2%) | 2 (2.4%) | 3 (1.3%) |
| Student | 3 (4.5%) | 2 (2.4%) | 3 (3.6%) | 8 (3.4%) |
| Volunteer | 1 (1.5%) | 0 (0.0%) | 0 (0.0%) | 1 (0.4%) |
| Unemployed or laid off | 5 (7.5%) | 8 (9.5%) | 11 (13.1%) | 24 (10.2%) |
| Unable to work | 4 (6.0%) | 10 (11.9%) | 13 (15.5%) | 27 (11.5%) |
| Missing or prefer not to answer | 2 (2.9%) | 5 (6.0%) | 2 (2.4%) | 9 (3.8%) |
| **Approximate annual household income** |  |  |  |  |
| Less than \$10,000 | 7 (10.4%) | 12 (14.3%) | 12 (14.3%) | 31 (13.2%) |
| \$10,000 to \$20,000 | 12 (17.9%) | 12 (14.3%) | 11 (13.1%) | 35 (14.9%) |
| \$20,000 to \$50,000 | 21 (31.3%) | 19 (22.6%) | 31 (36.9%) | 71 (30.2%) |
| \$50,000 to \$90, 000 | 9 (13.4%) | 14 (16.7%) | 11 (13.1%) | 34 (14.5%) |
| Greater than \$90,000 | 8 (11.9%) | 8 (9.5%) | 4 (4.8%) | 20 (8.5%) |
| Missing or prefer not to answer | 11 (16.2%) | 19 (22.6%) | 15 (17.9%) | 45 (19.1%) |
| **Insurance status<sup>1</sup>** |  |  |  |  |
| Medicare | 11 (16.2%) | 17 (20.2%) | 17 (20.2%) | 45 (19.1%) |
| Medicaid | 9 (13.2%) | 14 (16.7%) | 19 (22.6%) | 42 (17.8%) |
| Private insurance | 30 (44.1%) | 36 (42.9%) | 32 (38.1%) | 98 (41.5%) |
| Worker's compensation | 2 (2.9%) | 0 (0.0%) | 1 (1.2%) | 3 (1.3%) |
| Disability | 1 (1.5%) | 1 (1.2%) | 3 (3.6%) | 5 (2.1%) |
| No health insurance or missing | 19 (27.9%) | 20 (23.8%) | 20 (23.8%) | 59 (25.0%) |
| Prefer not to answer | 2 (2.9%) | 4 (4.8%) | 7 (8.3%) | 13 (5.5%) |

<sup>1</sup> Subjects were able to report more than one insurance type.

Baseline pain and clinical characteristics for the AA+UC, PA+UC, and UC groups are shown in **Table 2**. Overall, the most common primary pain locations were lower back (36.9%), legs (26.7%), and neck (14.4%). Most subjects (66.9%) reported having pain in more than one location, and 56.0% reported that their current painful condition was due to trauma or injury. The majority of subjects reported having at least some pain in the past 3 months (79.3%), and that pain had limited their life or work activities (66.5%). All three arms had similar ED baseline pain scores (AA+UC 7.0, SD 2.3, PA+UC 7.2, SD 2.2, UC 7.0, SD 2.1). Only 18% of study participants had ever tried acupuncture before. There were no differences in pre-baseline ED administration of opioid and non-opioid analgesics between the groups (**Table 2**).

**Table 2:**
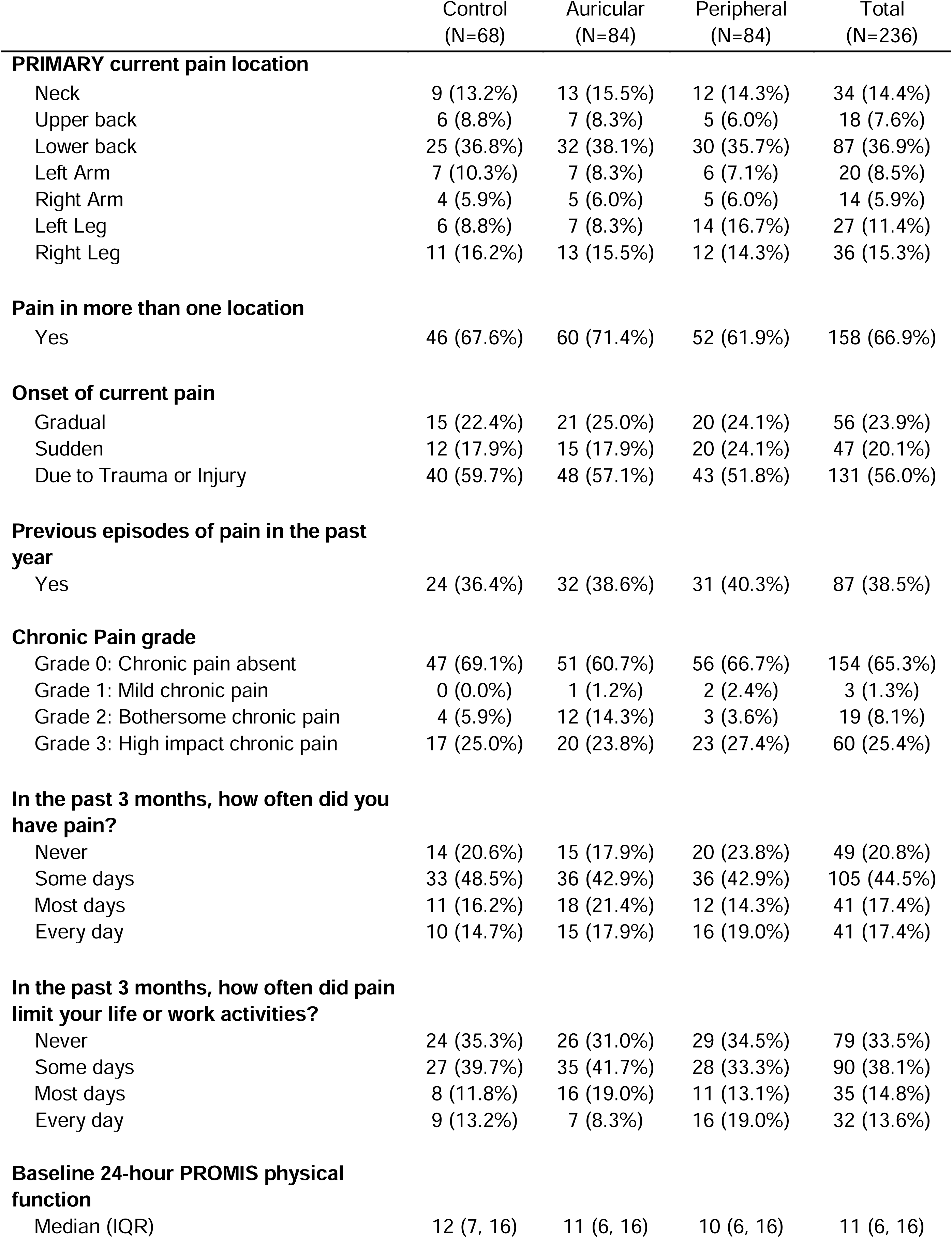
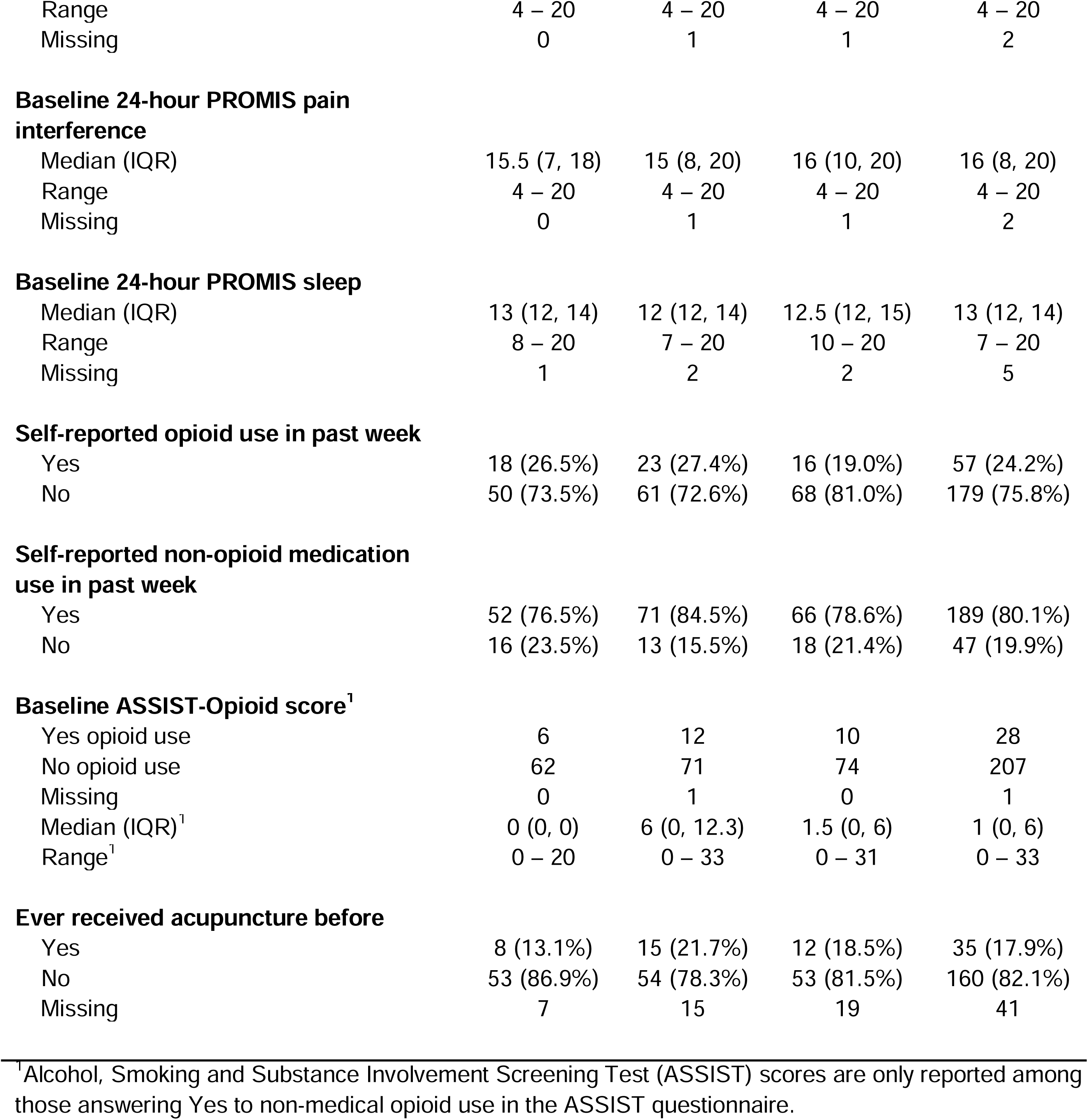
Baseline Clinical Characteristics.

### Main Results

**Table 3** shows the estimated reductions in pain at 1-hour post-treatment based on the multiple imputation model. Greater pain reductions at 1-hour were observed for both AA+UC (2.1; 95% CI: 1.6, 2.6) and PA+UC (1.6; 95% CI: 1.1, 2.1) compared with UC (0.5; 95% CI: -0.1, 1.0). Pain reductions with AA+UC and PA+UC did not differ significantly from each other. Although not significant, there were fewer opioids ordered in the ED or prescribed on ED discharge among the acupuncture groups compared to control at ED discharge (**Table 3**). Participants in both the AA+UC and PA+UC groups reported comparably high satisfaction with their overall acupuncture experience at 1-hr with a mean score of 4.4 (±0.9) on a 1-5 Likert scale (**Table 3**), demonstrating acceptability of the intervention.

**Table 3:** Pain Outcomes at 1 Hour.

|  | Control<br>(N=68) | Auricular<br>(N=84) | Peripheral<br>(N=84) | Total<br>(N=236) | P-Value |
| --- | --- | --- | --- | --- | --- |
| **Current pain NRS at time of triage** |  |  |  |  |  |
| Mean (SD) | 7.8 (2.0) | 7.7 (2.2) | 8.0 (2.1) | 7.8 (2.1) |  |
| Range | 3 – 10 | 2 – 10 | 2 – 10 | 2 – 10 |  |
| Missing | 2 | 0 | 1 | 3 |  |
| **Current pain NRS at study enrollment**<br>**(ED baseline)** |  |  |  |  |  |
| Mean (SD) | 7.0 (2.1) | 7.0 (2.3) | 7.2 (2.2) | 7.0 (2.2) |  |
| Range | (2-10) | (2-10) | (1-10) | (1-10) |  |
| Missing | 0 | 0 | 0 | 0 |  |
| **Current pain NRS at 1 hour** |  |  |  |  |  |
| Mean (SD) | 6.6 (1.9) | 4.9 (2.9) | 5.6 (2.6) | 5.6 (2.6) |  |
| Range | 2 – 10 | 0 – 10 | 0 – 10 | 0 – 10 |  |
| Missing | 4 | 3 | 2 | 9 |  |
| **Change in pain: triage to baseline**<br>**(enrollment)** |  |  |  |  |  |
| Mean (SD) | -0.8 (1.7) | -0.8 (1.8) | -0.7 (2.1) | -0.8 (1.9) |  |
| Range | (-8.0-2.0) | (-8.0-4.0) | (-9.0-7.0) | (-9.0-7.0) |  |
| 95% CI for mean | (-1.2, -0.4) | (-1.2, -0.3) | (-1.2, -0.3) | (-1.0, -0.5) |  |
| **Change in pain NRS at 1 hour** |  |  |  |  | <0.001<sup>1</sup> |
| Mean (SD) | -0.5 (2.0) | -2.1 (2.9) | -1.6 (1.9) | -1.5 (2.4) |  |
| Range | -6 – 5 | -9 – 7 | -7 – 5 | -9 – 7 |  |
| 95% CI for mean* | (-1.0, 0.1) | (-2.6, -1.6) | (-2.1, -1.1) | (-1.8, -1.1) |  |
| **Control – Auricular: Mean (95% CI)\*\*** |  | 1.6 (0.7, 2.6) |  |  |  |
| **Control – Peripheral: Mean (95% CI)\*\*** |  |  | 1.2 (0.3, 2.1) |  |  |
| **Percent change in pain at 1 hour** |  |  |  |  | 0.002<sup>2</sup> |
| Mean (SD) | 1.5 (50.7) | -23.8 (59.1) | -23.8 (33.7) | -16.9 (49.7) |  |
| Range | -95.3 – 250.0 | -100.0 – 350.0 | -100.0 – 125.0 | -100.0 – 350.0 |  |
| 95% CI for mean* | (-10.4, 13.5) | (-35.1, -12.6) | (-34.4, -13.2) | (-23.4, -9.7) |  |
| **Control – Auricular: Mean (95% CI)\*\*** |  | 25.4 (5.9, 44.9) |  |  |  |
| **Control – Peripheral: Mean (95% CI)\*\*** |  |  | 25.4 (6.0, 44.7) |  |  |
| **Difference in changes: (baseline to one-hour) - (triage to baseline)** |  |  |  |  | 0.005<sup>3</sup> |
| Mean (SD) | 0.4 (3.1) | -1.4 (4.0) | -0.9 (2.9) | -0.7 (3.5) |  |
| Range | (-4.0-13.0) | (-11.0-8.0) | (-12.0-8.0) | (-12.0-13.0) |  |
| 95% CI for mean | (-0.4, 1.2) | (-2.3, -0.5) | (-1.6, -0.3) | (-1.2, -0.3) |  |
| Missing | 6 | 3 | 2 | 11 |  |
| **ED Analgesia Received Pre-Baseline** | 37 (54.4%) | 54 (64.3%) | 46 (54.8%) | 137 (58.1%) |  |
| Opioid pre-baseline | 17 (25.0%) | 22 (26.2%) | 17 (20.2%) | 56 (23.7%) |  |
| Non-opioid pre-baseline | 31 (45.6%) | 49 (58.3%) | 40 (47.6%) | 120 (50.8%) |  |
| **Analgesia Post-Baseline to 1 hour** | 24 (35.3%) | 22 (26.2%) | 17 (20.2%) | 63 (26.7%) | 0.113<sup>4</sup> |
| Opioid post-Baseline to 1 hour | 9 (13.2%) | 6 (7.1%) | 5 (6.0%) | 20 (8.5%) | 0.238<sup>4</sup> |
| Non-opioid post-baseline to 1 hour | 21 (30.9%) | 17 (20.2%) | 13 (15.5%) | 51 (21.6%) | 0.067<sup>4</sup> |
| **Discharge opioid prescription** | 10 (14.7%) | 8 (9.5%) | 6 (7.1%) | 24 (10.2%) | 0.169<sup>4</sup> |
| **Overall satisfaction with acupuncture at 1 hour** |  |  |  |  | 0.51<sup>4</sup> |
| Mean (SD) |  | 4.3 (1.0) | 4.4 (0.9) | 4.4 (0.9) |  |
| Range |  | (1.0-5.0) | (1.0-5.0) | (1.0-5.0) |  |
| 95% CI for mean |  | (4.1, 4.5) | (4.2, 4.6) | (4.2, 4.5) |  |
| Missing | NA | 7 | 9 | 16 |  |

Missing values were imputed for 1-hour change score calculations. NA=Not Applicable.
<sup>1</sup> p-value from overall ANOVA test. P-values for pairwise comparisons, adjusting for multiple comparisons using Bonferroni's method, are:
Control vs Auricular: <0.001
Control vs Peripheral: 0.002
Auricular vs Peripheral: 0.21
<sup>2</sup> p-value from overall ANOVA test. P-values for pairwise comparisons, adjusting for multiple comparisons using Bonferroni's method, are:
Control vs Auricular: 0.002
Auricular vs Peripheral: 1.00
<sup>3</sup> ANOVA overall F-Test. P-values for pairwise comparisons, adjusting for multiple comparisons using Tukey's method, are:

**Table 3** shows the mean pain score at triage, baseline and 1-hr post-treatment, as well as the multiple-imputation based estimates of change from baseline to 1-hr post-treatment. Changes in pain score from ED arrival at triage to pre-intervention study baseline were similar across the groups, with the overall sample mean change in pain score of -0.8 (± 1.9). Moreover, similar numbers of pain medications were given during this time interval among the three groups, indicating a similar effect of usual care ED medications at baseline. The AA+UC and PA+UC groups had significant decreases in pain at 1-hour post-treatment while the UC group did not. For the exploratory analyses, the AA+UC group showed significantly better within-group improvements after acupuncture than after usual care alone, while the PA+UC group showed a similar trend that was not significant. Participants in both acupuncture groups also received fewer opioid and non-opioid analgesics post-baseline and at discharge than control, although these findings were not statistically significant. There was no impact of pre-existing chronic pain on pain response for any treatment arm nor as a main effect in the exploratory models. The results of the complete case analyses were similar to those of the ITT analyses (**eTable1**).

Overall, there were few AEs and no serious adverse events (SAEs) reported (**eTable2**). There was no significant difference in the number of subjects who reported AEs in the UC (n=2, 2.9%), AA+UC (n=3, 3.6%), and PA+UC (n=0, 0.0%) groups in the ED. The most common AEs were transient pain, bleeding or bruising at needle sites, self-limited headache or brief episodes of anxiety.

## LIMITATIONS

Limitations of this study include a single urban ED in southeastern U.S. which may limit generalizability to other environments, such as rural ED’s and other geographic locations. However, studies from other countries have shown similar efficacy of ED acupuncture for treating pain, and ours adds to this body of literature as one of the largest U.S.-based studies. Enrollment was limited to English-speaking patients due to a lack of validated non-English versions for most of the questionnaires used, which may limit applicability to non-English speaking populations. However, only 2.3% of screen fails were due to not speaking English and our final study population reflects the population seen in this setting, including the distribution of Hispanic ethnicity among our patients. Treatment delivery by licensed acupuncturists may limit comparison to other ED-based trial protocols that trained other providers, like physicians, or environments with no available acupuncturists. While every attempt was made to blind ED providers and outcomes assessors, patients could not be blinded to treatment assignment. However, we accounted for potential placebo effects of extra clinician time and attention by having study acupuncturists evaluate and interact with all control participants. Future research should include multi-site RCT’s with varied ED settings to further evaluate acupuncture’s efficacy across patient groups and practice environments.

## DISCUSSION

### Improved acute musculoskeletal pain management

Effective management of acute pain is critically important to mitigate associated morbidity and disability; however, the current reliance on opioid medications presents substantial risks.^8,42^ Previous studies of acupuncture in the ED have shown greater improvements than sham acupuncture and similar benefits as medications for treating acute pain, but have been limited by small sample sizes.^13,22,26,43^ Our study builds upon prior work with a uniquely large and diverse population of ED patients in a pragmatic randomized controlled trial design. Our study shows clinically significant improvements in pain scores 1-hour after acupuncture compared to usual care. We found that two different acupuncture interventions were similarly effective, allowing for increased flexibility in terms of both patient preference and clinical implementation.

There were fewer ED opioid administrations and discharge prescriptions after acupuncture. While this finding was not statistically significant, many pain experts agree that any prevention of new opioid use is clinically meaningful as it prevents the negative sequelae of opioid side effects and misuse.^44^ Moreover, pain improvements were better with acupuncture, addressing the concern that reduction in opioids may have led to undertreatment of pain. Previous work in ED and cancer patients has shown that acupuncture can outperform opioids in treating pain and may reduce opioid prescriptions.^21,45,46^ One study, comparing acupuncture and intravenous morphine for acute pain in an ED setting, found that acupuncture was more likely to cause significant reduction in pain (≥50%), and faster, than morphine alone.^21^ Another study comparing acupuncture patients with those using non-steroidal anti-inflammatory drugs (NSAIDs) and physical therapy found that acupuncture patients were prescribed fewer opioids and had fewer ED visits.^46^ Thus, acupuncture may be an important treatment option for reducing opioid prescribing and subsequent use.

### Delivery of acupuncture to a broad population in the ED environment

Ours is one of the first pragmatic randomized controlled trials of acupuncture to intentionally and successfully enroll a large number of people from underserved and minoritized groups in the U.S.^16,47,48^ By minimizing the number of exclusion criteria that have historically excluded these populations,^49^ and systematically approaching all potentially qualifying patients for the study, our study population is reflective of our general ED population.^50^ More than 50% of participants self-identified as Black and 7% as Latino, more than half reported low income <$50,000, and over half had public or no health insurance. These rates are higher than U.S. national averages reporting 36.1% with income under $50,000^51^ and 8.3% with no health insurance.^52^ Few prior studies of acupuncture report the race of participants. Ours is the first randomized trial to our knowledge to demonstrate acupuncture efficacy across a diverse and underrepresented population.

Furthermore, we successfully adapted acupuncture to a fast-paced, relatively chaotic ED environment by keeping treatments between 20 to 30 minutes and focusing on pain relief and needling of sites easily accessible while seated in a chair or laying in a stretcher fully clothed.^53,54^ Our ability to recruit and perform acupuncture on 236 ED patients over a one-year period demonstrates feasibility. Participants in both acupuncture intervention arms reported high patient satisfaction and minimal side effects, demonstrating broad acceptability across diverse populations. Our findings underscore those from recent work developing community acupuncture clinics for underserved populations reporting high participant interest in and satisfaction with acupuncture treatments,^18,47,55,56^ supporting implementation of acupuncture more broadly to a wide range of patients.

## CONCLUSION

These results indicate that both auricular and peripheral acupuncture are feasible, acceptable, and effective in the emergency department for acute musculoskeletal pain and should be further explored for more widespread implementation.

## Data Availability

Data sharing is not currently available due to ongoing additional analyses. Data available in the future on request.

## ACKNOWLEDGEMENTS

We would like to acknowledge all of the members of the **Duke Emergency Department Acupuncture Research** team, including licensed acupuncturists Christi De Larco, Michelle Mill, Austin Dixon and Tara Bianca Rado; clinical research coordinators Erica Walker, Tedra Porter, Andrew Bouffler, Lauren McGowan, and Madison Frazier; research assistants Morgan Seifert and Sophie Finkelstein; and Duke Integrative Medicine for their integral partnership on this project.

## Copyright

The Corresponding Author has the right to grant on behalf of all authors and does grant on behalf of all authors, an exclusive license on a worldwide basis to the American College of Emergency Physicians (ACEP) to permit this article (if accepted) to be published in *Annals of EM* editions and any other ACEP products and sublicences such use and exploit all subsidiary rights, as set out in our license.

## Ethics approval

The Duke University Health System Institutional Review Board has reviewed and approved this study on January 29, 2020 (Protocol No: Pro00104140).

## Clinicaltrials.gov registration

Emergency Department Acupuncture for Acute Musculoskeletal Pain Management ID#: NCT04290741; URL: https://clinicaltrials.gov/ct2/show/NCT04290741

---

**eTable1:** Updated Secondary Analysis: One-Hour Change-in-Pain.

|  | 1. Control<br>(N=68) | 2. Auricular<br>(N=84) | 3. Peripheral<br>(N=84) | Total<br>(N=236) | p value |
| --- | --- | --- | --- | --- | --- |
| **Change in pain at 1 hour** |  |  |  |  | <0.001<sup>1</sup> |
| Missing | 4 | 3 | 2 | 9 |  |
| Mean (SD) | -0.3 (2.0) | -2.2 (2.9) | -1.7 (.9) | -1.5 (2.4) |  |
| Median | 0.0 | -2.0 | -1.0 | -1.0 |  |
| Q1, Q3 | -1.0, 0.0 | -4.0, 0.0 | -3.0, 0.0 | -3.0, 0.0 |  |
| Range | (-5-5) | (-9-7) | (-7-5) | (-9-7) |  |
| **Percentage change in pain at 1 hour** |  |  |  |  | <0.001<sup>2</sup> |
| Missing | 4 | 3 | 2 | 9 |  |
| Mean (SD) | 3.7 (51.3) | -25.6 (57.5) | -24.4 (33.6) | -16.9 (49.7) |  |
| Median | 0.0 | -28.6 | -21.1 | -16.7 |  |
| Q1, Q3 | -16.7, 0.0 | -60.0, 0.0 | -40.0, 0.0 | -40.0, 0.0 |  |
| Range | (-71.4-250.0) | (-100.0-350.0) | (-100.0-125.0) | (-100.0-350.0) |  |

<sup>1</sup> p-value from overall ANOVA test. P-values for pairwise comparisons, adjusting for multiple comparisons using Tukey's method, are:
Control vs Auricular: <0.001
Control vs Peripheral: 0.003
Auricular vs Peripheral: 0.34
<sup>2</sup> p-value from overall ANOVA test. P-values for pairwise comparisons, adjusting for multiple comparisons using Tukey's method, are:
Control vs Auricular: 0.001
Control vs Peripheral: 0.002
Auricular vs Peripheral: 0.98

**eTable2:** Adverse Events (AEs). Patient-reported AEs in the ED included transient needle site pain or irritation (n=3), and non-acupuncture related episodes of fainting related to blood draw (n=1) and nausea related to opioid medication (n=1), respectively, as part of usual ED care. Some patients reported more than one adverse event.

| **eTable2: Adverse Events** |  |  |  |  |  |
| --- | --- | --- | --- | --- | --- |
|  | Control<br>(N=68) | Auricular<br>(N=84) | Peripheral<br>(N=84) | Total<br>(N=236) | p value |
| **Number of AEs in ED** |  |  |  |  | 0.37<sup>1</sup> |
| 0 | 66 (97.1%) | 81 (96.4%) | 84 (100.0%) | 231 (97.9%) |  |
| 1 | 2 (2.9%) | 2 (2.4%) | 0 (0.0%) | 4 (1.7%) |  |
| 3 | 0 (0.0%) | 1 (1.2%) | 0 (0.0%) | 1 (0.4%) |  |
| <sup>1</sup>Fisher Exact |  |  |  |  |  |

